# Non-Motor symptoms in prodromal Parkinson’s disease are linked to reduced Quality of Life

**DOI:** 10.1101/2023.09.19.23295779

**Authors:** Sinah Röttgen, Marie-Sophie Lindner, Aline Seger, Johanna Kickartz, Kim-Lara Weiß, Christopher E J Doppler, Gereon R Fink, Anja Ophey, Michael Sommerauer

**Author notes:** Corresponding author: Dr. Michael Sommerauer, Department of Neurology, Faculty of Medicine and University Hospital Cologne, University of Cologne, Kerpener Straße 62, 50937 Köln, Germany;, phone: +49 221/478 −6191.

## Abstract

**Background:** Isolated REM sleep behavior disorder (iRBD) is the most specific prodromal marker of Parkinson’s disease (PD), often accompanied by various non-motor symptoms. In PD, non-motor symptoms are strongly associated with reduced health-related quality of life (hrQoL).

**Objectives:** To identify factors linked to poorer hrQoL in patients with iRBD and to compare their hrQoL to healthy control participants (HC) and patients with PD.

**Methods:** Sixty-two patients with iRBD (*M* = 66.44±6.14), 29 patients with PD (*M* = 67.21±7.23), and 19 HC (*M* = 67.57±8.16) were included. We administered the 36-Item Short Form Health Survey (SF-36) to assess hrQoL. Additionally, participants underwent a comprehensive clinical evaluation of non-motor symptoms.

**Results:** The SF-36 total score was significantly lower in patients with iRBD (83.33±16.96) compared to HC (92.29±5.49, U = 390.00, Z = −2.218, p = .027, r = 0.246). Poorer hrQoL in patients with iRBD was linked to self-reported neuropsychiatric symptoms, sleep-wake disturbances, and a higher burden of autonomic symptoms (all *r* = -.25 to -.76, all *p* < .05). Multiple regression analysis revealed fatigue and depressive symptoms as significant predictors of poorer hrQoL in patients with iRBD (*F* (5.56) = 51.59, *p* < .001, adjusted *R*^2^ = 0.81).

**Conclusions:** This study highlights the importance of non-motor symptoms for hrQoL in prodromal PD, irrespective of motor symptoms. Fatigue and depressive symptoms arise as the most relevant therapeutic targets in the prodromal stage of PD to optimize the patient’s quality of life.

## 1 Introduction

Isolated rapid eye movement (REM) sleep behavior disorder (iRBD) is a parasomnia defined by dream-enacting behavior and a loss of muscle atonia during REM sleep^1,2^. Compelling evidence identified iRBD as an incipient α-synucleinopathy^3–5^. To date, iRBD is the most specific prodromal marker of Parkinson’s disease (PD) and can precede the onset of PD-defining motor symptoms up to decades^6,7^.

By definition, patients with iRBD do not exhibit severe motor symptoms, but mild motor disturbances can be observed^8–10^. In contrast, non-motor symptoms such as constipation, olfactory dysfunction, and depressive symptoms are frequent in iRBD^11^. A recent systematic review and meta-analysis reported a prevalence of depression in patients with iRBD of 28.8% and an odds ratio for depression of 2.57^12^, indicating disturbed mental well-being in prodromal PD.

Health-related Quality of Life (hrQoL) is increasingly recognized as the essential factor for the impact of a disease on a patient’s life. In PD, a significantly lower hrQoL compared to HC has been reported^13^. Interestingly, the burden of non-motor symptoms in PD, particularly depressive symptoms, showed a stronger association with hrQoL than motor symptoms^13,14^. Data on hrQoL in iRBD are scarce, and only one study assessed hrQoL in iRBD, showing reduced hrQoL in iRBD patients compared to HC^15^. Moreover, identifying factors associated with reduced hrQoL is vital, particularly in prodromal PD, as it may help develop therapeutic interventions to restore hrQoL without disease-modifying agents.

It is, therefore, of broad clinical interest to understand which symptoms arising in the prodromal stage of PD are linked to hrQoL. In the present study, we studied hrQoL in patients with iRBD compared to HC and patients with PD, and its relation to emerging motor symptoms and a variety of non-motor symptoms. We hypothesized that hrQoL is diminished in patients with iRBD compared to HC but still better than in PD. Comparable to PD, non-motor symptoms may contribute to poorer hrQoL in patients with iRBD beyond demographic variables and subtle motor symptoms.

## 2 Materials and Methods

### 2.1 Study design and participants

Participants were recruited from July 2020 to November 2021 at the University Hospital of Cologne from our ongoing iRBD discovery cohort^16^. Inclusion criteria for patients with iRBD were age 50 to 80 years, the ability to give informed consent, and polysomnography-confirmed iRBD according to the International Classification of Sleep Disorders III criteria^1,16^. The exclusion criterion was a significant cognitive impairment operationalized by the Montreal Cognitive Assessment (MoCA; cutoff <22)^17^. We also included patients with PD diagnosed according to the current consensus criteria^18^ with identical in- and exclusion criteria. For HC, depressive symptoms operationalized by the Beck’s Depression Inventory II (BDI-II; cutoff ≥9)^19^ and symptoms of RBD or any movement disorder were additional exclusion criteria. Patients with PD and HC were recruited from other studies of the investigators.

All subjects gave written informed consent prior to participation. The local ethics committee of the Medical Faculty of the University of Cologne approved the study.

### 2.2 Assessment of health-related quality of life

HrQoL was measured with the 36-Item Short Form Health Survey (SF-36)^20^, a cross-disease patient-reported health status survey. A systematic review including 29 studies showed that the SF-36 questionnaire is the most frequently used generic assessment to estimate hrQoL in PD^13^. As the SF-36 is not disease-specific, it is better suited to investigate hrQoL in the prodromal stage of PD than PD-specific hrQoL questionnaires, which typically focus on motor-related symptom burden. The questionnaire consists of 36 items assessing eight hrQoL domains: physical function, role limitations due to physical problems (physical role), bodily pain, general health, vitality, social functioning, role limitations due to emotional problems (emotional role), and psychological well-being. Most questions use Likert scaling, but several subitems have a dichotomous (yes/no) format. The questionnaire is scored by recoding item responses first and subsequently averaging items of one domain to create the eight SF-36 domain scores. The total score indicates the arithmetic average of all domain scores. Domain scores and the total score range from 0 to 100, with higher scores indicating better hrQoL.

### 2.3 Clinical Assessment

Subjects completed questionnaires on various non-motor symptoms. Sleep quality was evaluated by the Pittsburgh Sleep Quality Index (PSQI)^21^. Cognitive, emotional, and behavioral features of psychophysiological insomnia were measured by the Regensburg Insomnia Scale (RIS)^22^. The Epworth Sleepiness Scale (ESS)^23^ interrogates daytime sleepiness. Presence and severity of anxiety were determined using Beck’s Anxiety Inventory (BAI)^24^, and depressive symptoms were assessed with the BDI-II^25^. Apathy was assessed by the Apathy Evaluation Scale (AES)^26^, and fatigue was examined by the Fatigue Scale for Motor and Cognitive Functions (FSMC)^27^. Subjective cognitive impairment (SCI) was measured by an extended version of the Subjective Memory Impairment Questionnaire (SMI-Q)^28,29^. We assessed the SCOPA-AUT^30^ questionnaire to examine autonomic symptoms and the non-motor symptom questionnaire (NMSQ)^31^ for non-motor symptoms in general. For all questionnaires, higher total scores indicate a higher symptom burden.

All participants underwent a comprehensive clinical work-up including the 12-item Sniffin’ Sticks test (Burkhardt®, Wedel, Germany), cognitive screening with the MoCA^17^, and motor symptoms were quantified with part III of the MDS-UPDRS^32^ (except for HC participants).

### 2.4 Statistical analyses

Statistical analyses were conducted using SPSS version 28.0^33^ and R Studio^34^. The normality of data was assessed with Shapiro-Wilk and Kolmogorov-Smirnov tests and Q-Q plots. As all data were not normally distributed. The Kruskall-Wallis test was used to assess group differences (iRBD, HC, PD). Post-hoc tests were performed using Mann-Whitney-U tests. A chi-Square test was performed to interrogate categorical variables. The relationship between hrQoL and non-motor symptoms and clinical and demographic data was analyzed with Spearman partial correlations, including age and sex as covariates. Based on a significant correlation in the bivariate partial correlation analyses (*p* < .05), non-motor symptom scales were selected as potential determinants of hrQoL in multiple linear regression analyses for the subgroup of patients with iRBD. PSQI, RIS, ESS, BAI, BDI-II, AES, FSMC, SCI, SOPA-AUT, and NMSQ were included as independent variables in a stepwise regression approach. The SF-36 total score and the eight SF-36 domain scores were used as dependent variables. All regression models included age and sex as covariates. The significance level was set at *p* < .05 uncorrected.

### 2.5 Data Availability Statement

The data included in this study are available on reasonable request to the corresponding author.

## 3 Results

### Sample Characteristics

The analyses included sixty-two patients with iRBD, 19 HC participants, and 29 PD patients. Patients with iRBD were comparable concerning age and sex to both other groups. Patients with iRBD were aged 54 to 78 years (66.44 ± 6.14), 54 of whom were male (87%), and none reported any relevant motor impairment during daily routine. Demographic and clinical characteristics are shown in Table 1.

**Table 1.** Demographic and clinical characteristics of included participants.

|  | HC<br>participants<br>(n=19) | iRBD<br>patients<br>(n=62) | PD<br>patients<br>(n=29) | p<br>all<br>groups | Post-hoc<br>tests |
| --- | --- | --- | --- | --- | --- |
| Gender (male/female) | 14/5 | 54/8 | 22/7 | .260<sup>#</sup> | (HC = iRBD = PD) |
| Age [years] | 67.57 ±8.16 | 66.44 ±6.14 | 67.21 ±7.23 | .806 | (HC = iRBD = PD) |
| MoCA | 26.53 ±1.84 | 27.50 ±1.65 | 26.55 ±2.28 | .036 | ((iRBD = PD) > HC) |
| Sniffin' Sticks | 9.79 ±1.55 | 6.32 ±2.76 | 5.17 ±2.56 | ≤.001 | (HC > iRBD > PD) |
| MDS-UPDRS-III OFF | NA | 3.92 ±2.56 | 38.48 ±16.1 | ≤.001 | (iRBD < PD) |
| Disease duration [years]* | NA | 7.58 ±6.17 | 6.78 ±4.93 |  |  |
| **SF-36 assessment** |  |  |  |  |  |
| Body function | 96.58 ±5.28 | 92.26 ±10.58 | 71.38 ±22.91 | ≤.001 | ((HC = iRBD) > PD) |
| Physical role | 96.05 ±9.37 | 84.27 ±32.4 | 47.41 ±41.91 | ≤.001 | ((HC = iRBD) > PD) |
| Bodily pain | 88.05 ±19.09 | 80.97 ±27.01 | 63.48 ±28.48 | .002 | ((HC = iRBD) > PD) |
| General health | 85.05 ±9.71 | 72.74 ±18.05 | 48.45 ±24.18 | ≤.001 | (HC > iRBD > PD) |
| Vitality | 85.26 ±8.58 | 73.95 ±20.17 | 59.83 ±16.39 | ≤.001 | (HC > iRBD > PD) |
| Social functioning | 96.05 ±11.82 | 91.53 ±17.5 | 78.88 ±18.33 | ≤.001 | ((HC = iRBD) > PD) |
| Emotional role | 98.25 ±7.65 | 88.17 ±30.83 | 72.41 ±37.87 | .004 | ((HC = iRBD) > PD) |
| Psychological well-being | 93.05 ±4.96 | 82.77 ±16.96 | 71.86 ±16.37 | ≤.001 | (HC > iRBD > PD) |
| Total score | 92.29 ±5.49 | 83.33 ±16.96 | 64.21 ±17.48 | ≤.001 | (HC > iRBD > PD) |
| **Non-motor symptoms** |  |  |  |  |  |
| PSQI | 4.16 ±2.54 | 4.73 ±3.11 | 7.50 ±3.86 | ≤.001 | ((HC = iRBD) < PD) |
| RIS | 6.58 ±4.95 | 6.95 ±5.34 | 11.67 ±5.65 | ≤.001 | ((HC = iRBD) < PD) |
| ESS | 6.21 ±3.07 | 5.53 ±3.41 | 9.38 ±3.72 | ≤.001 | ((HC = iRBD) < PD) |
| BAI | 1.05 ±1.47 | 3.82 ±5.32 | 8.97 ±7.86 | ≤.001 | (HC < iRBD < PD) |
| BDI | 1.37 ±1.71 | 5.71 ±7.26 | 8.69 ±6.76 | ≤.001 | (HC < iRBD < PD) |
| AES | 21.84 ±3.13 | 28.00 ±8.14 | 28.38 ±8.10 | .007 | (HC < (iRBD = PD)) |
| FSMC | 25.58 ±5.94 | 30.85 ±13.24 | 45.90 ±16.49 | ≤.001 | ((HC = iRBD) < PD) |
| SCI | 3.16 ±3.24 | 6.45 ±5.54 | 10.17 ±8.37 | ≤.001 | (HC < iRBD < PD) |
| SCOPA-AUT | 4.26 ±3.28 | 6.94 ±3.56 | 13.52 ±7.50 | ≤.001 | (HC < iRBD < PD) |
| NMSQ | 1.53 ±1.50 | 5.37 ±3.73 | 8.43 ±4.67 | ≤.001 | (HC < iRBD < PD) |

*Note.* Results are expressed as mean ± standard deviation or numbers. Groups were compared using Kruskal-Wallis tests. Post-hoc comparisons were performed with Mann-Whitney U tests. <sup>#</sup>Chi-Square test. \*The disease duration for patients with iRBD is the time of self-reported iRBD symptoms in years, and for patients with PD, the time in years since PD diagnosis. HC = healthy controls; iRBD = isolated REM sleep behavior disorder; PD = Parkinson's disease; MoCA = Montreal Cognitive Assessment; MDS-UPDRS-III OFF = MDS-Unified Parkinson's Disease Rating Scale Part III during the OFF phase; SF-36 = Short Form Health Survey 36; PSQI = Pittsburgh Sleep Quality Index; RIS = Regensburg Insomnia Scale; ESS = Epworth Sleepiness Scale; BAI = Beck Anxiety Inventory; BDI = Beck Depression Inventory-II; AES = Apathy Evaluation Scale; FSMC = Fatigue Scale for Motor and Cognitive Functions; SCI = Subjective Cognitive Impairment; SCOPA-AUT = Scales for Outcomes in Parkinson's Disease - Autonomic questionnaire; NMSQ = Non-Motor Symptoms Questionnaire. No data on MDS-UPDRS-III off was available for control subjects.

Significant group differences were observed for PSQI (*H*(2) = 13.54, *p* = .001), RIS (*H*(2) = 14.07, *p* = .001), ESS (*H*(2) = 19.34, *p* < .001), and FSMC (*H*(2) = 25.95, *p* < .001). Post-hoc analysis revealed no significant differences between HC and iRBD. Nevertheless, both HC and iRBD exhibited significantly lower scores when compared to PD. Further significant group differences were observed for BAI (*H*(2) = 28.40, *p* < .001), BDI (*H*(2) = 22.78, *p* < .001), SCI (*H*(2) = 16.60, *p* < .001), SCOPA-AUT (*H*(2) = 27.87, *p* < .001), and NMSQ (*H*(2) = 34.77, *p* < .001). Post-hoc tests indicated significantly higher values in patients with iRBD than HC but lower than in PD. In addition, AES (*H*(2) = 9.82, *p* = .007) differed significantly between groups. Post-hoc tests showed no differences between iRBD and PD. However, both iRBD and PD displayed significantly higher scores when compared to HC.

### Health-related quality of life in iRBD compared to PD and HC

A Kruskal-Wallis test revealed significant group differences in the SF-36 total score (*H*(2) = 35.32, *p* < .001). HC participants had a significantly higher SF-36 total score than patients with iRBD (*U* = 390.00, *Z* = −2.218, *p* = .027, *r* = 0.246), but patients with PD had a significantly lower SF-36 total score compared to iRBD (*U* =323.50, *Z* = −4.902, *p* < .001, *r* = 0.514) and HC (*U* = 30.00, *Z* = −5.176, *p* < .001, *r* = 0.747). Furthermore, there were significant differences between patients with iRBD and HC in the SF-36 domains general health (*U* = 346.00, *Z* = −2.720, *p* = .007, *r* = 0.302), vitality (*U* = 402.00, *Z* = −2.096, *p* = .036, *r* = 0.233), and psychological well-being (*U* = 375.50, *Z* = −2.414, *p* = .016, *r* = 0.268, see Figure 1). HrQoL in PD was significantly lower than in patients with iRBD and HC in all SF-36 domains (all *U* = 37.00 to 673.50, all *Z* = −5.056 to 2.606, all *p* < .01, all *r* = 0.273 to 0.730).

**Figure 1.**
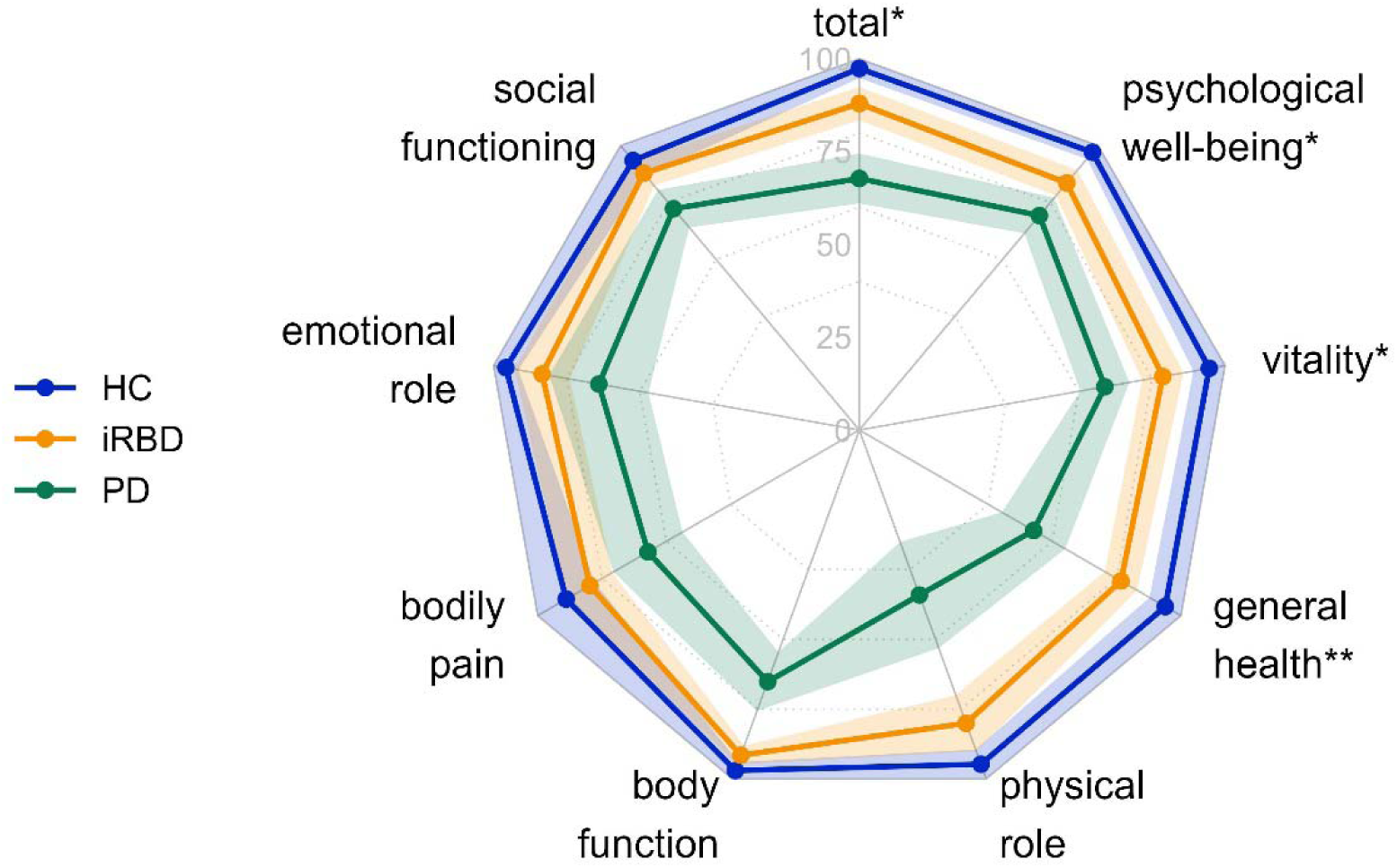
HrQoL comparisons of all SF-36 domains as well as the total score. *Note*. N = 110. Dots express mean values, and colored areas show 95% confidence intervals. HC = Healthy control participants (n = 19); iRBD = iRBD patients (n = 62); PD = PD patients (n = 29). SF-36 scores range from 0 to 100, with higher scores indicating better hrQoL. * p < .05, ** p < .01. Asterisks indicate significant group differences between HC and iRBD in the post-hoc Mann-Whitney-U tests. All group differences between iRBD and PD were significant (all p < .01).

### Factors associated with health-related quality of life in iRBD

To analyze relevant factors linked to HrQoL in patients with iRBD, we correlated HrQoL domains and the SF-36 total score with outcomes of the clinical and non-motor assessment. Most prominently, we observed a close link between pronounced neuropsychiatric symptoms and reduced hrQoL in various domains. When correcting for age and sex with partial correlation analysis, PSQI, RIS, ESS, BAI, BDI-II, AES, FMSC, SCI, SCOPA-AUT, and NMSQ scores were negatively correlated with hrQoL in patients with iRBD (all *r* = -.25 to -.76, *all p* < .05, see Figure 2). MoCA was positively correlated with vitality in patients with iRBD (*r* = .25, *p* = .039). Interestingly, incipient motor symptoms were not associated with hrQoL in patients with iRBD (all *r* = -.02 to -.12).

**Figure 2.**
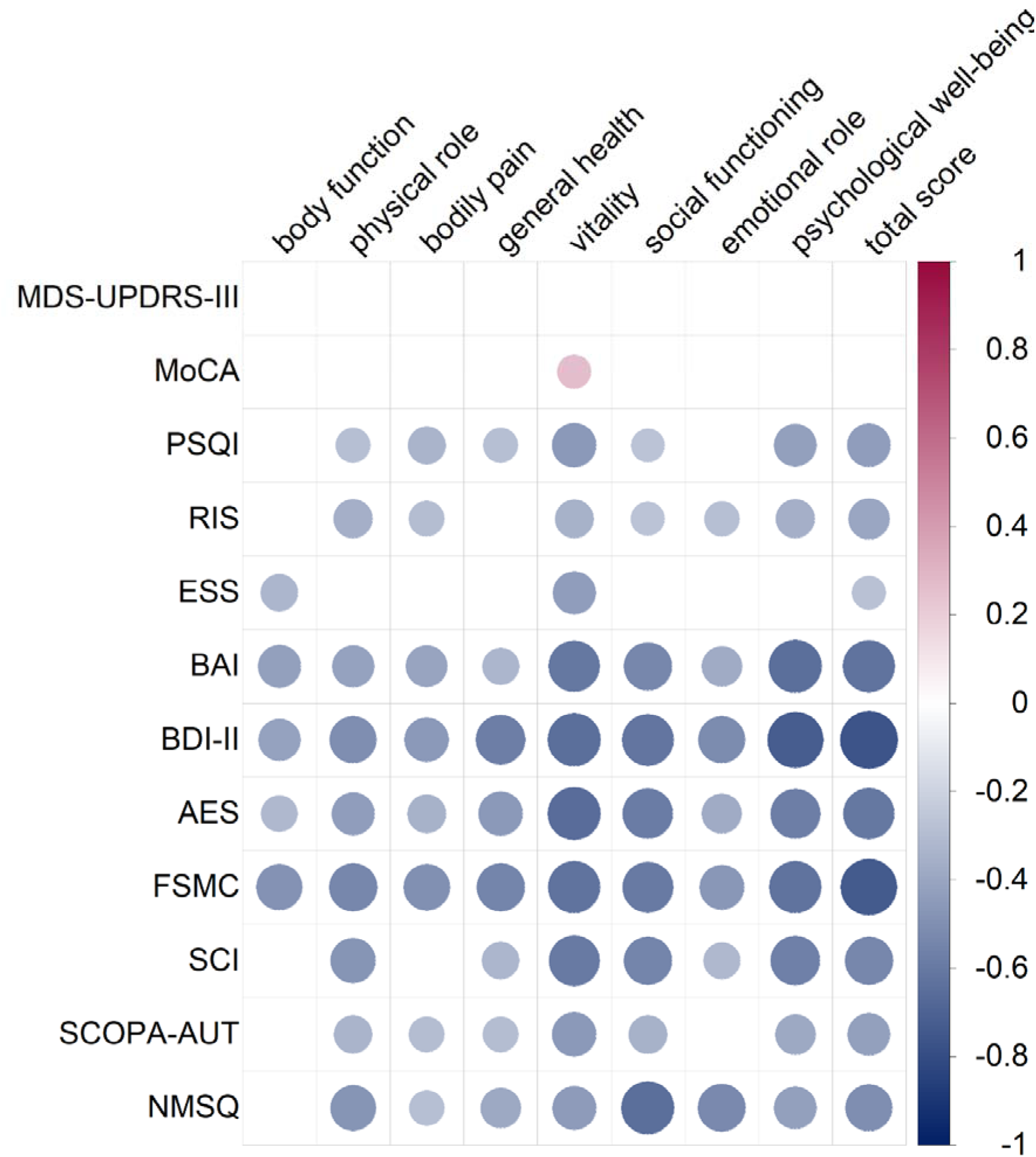
Partial correlation analyses (corrected for age and sex) of all SF-36 questionnaire domains and the total score with clinical variables and neuropsychiatric symptoms in iRBD patients. *Note.* N = 62. The significance level for all Spearman partial correlations was set to .05. Bigger dots indicate more significant correlations, with blue-colored dots showing negative correlations and red-colored dots illustrating positive correlations. MDS-UPDRS-III = MDS-Unified Parkinsońs Disease Rating Scale Part III; MoCA = Montreal Cognitive Assessment; PSQI = Pittsburgh Sleep Quality Index; RIS = Regensburg Insomnia Scale; ESS = Epworth Sleepiness Scale; BAI = Beck Anxiety Inventory; BDI-II = Beck Depression Inventory II; AES = Apathy Evaluation Scale; Fatigue = Fatigue Scale for Motor and Cognitive Functions; SCI = Subjective Cognitive Impairment; SCOPA-AUT = Scales for Outcomes in Parkinsońs Disease - Autonomic questionnaire; NMSQ = Non-Motor Symptoms Questionnaire.

Based on the significant bivariate correlation analyses (*p* < .05), PSQI, RIS, ESS, BAI, BDI-II, AES, FMSC, SCI, SCOPA-AUT, and NMSQ were selected as potential candidates for the multiple linear regression analyses. The adjusted *R*^2^ of the multiple regression analyses controlling for age and sex using a stepwise approach to evaluate the independent influence of the clinical scales ranged from .345 to .805. PSQI, RIS, AES, and NMSS were each a predictor of singular SF-36 domains: total score, body role, vitality, and emotional role, respectively (see Table 2). However, fatigue and depressive symptoms were significant determinants of a large set of hrQoL subdomains of the SF-36.

**Table 2.** Multiple regression analyses (corrected for age and sex) of all SF-36 domains and the total score in iRBD patients.

|  | PSQI | RIS | BDI | AES | FSMC | NMSQ | ANOVA | R<sup>2</sup> | Adj. R<sup>2</sup> |
| --- | --- | --- | --- | --- | --- | --- | --- | --- | --- |
| Total score | -0.143* | | -0.363*** | | -0.546*** | | $F(5.56) = 51.516$,<br>$p < .001$ | 0.821 | 0.805 |
| Body function | | | | | -0.585*** | | $F(3.58) = 12.964$,<br>$p < .001$ | 0.401 | 0.370 |
| Body role | | -0.204* | | | -0.691*** | | $F(4.57) = 20.710$,<br>$p < .001$ | 0.592 | 0.564 |
| Pain | | | | | -0.657*** | | $F(3.58) = 14.472$,<br>$p < .001$ | 0.428 | 0.399 |
| General health | | | -0.585*** | | | | $F(3.58) = 11.720$,<br>$p < .001$ | 0.377 | 0.345 |
| Vitality | | | -0.334* | -0.279** | -0.321** | | $F(5.56) = 25.877$,<br>$p < .001$ | 0.698 | 0.671 |
| Social functioning | | | -0.596*** | | -0.321** | | $F(4.57) = 39.547$,<br>$p < .001$ | 0.735 | 0.717 |
| Emotional role | | | | | -0.384** | -0.313* | $F(4.57) = 11.813$,<br>$p < .001$ | 0.453 | 0.415 |
| Psychological well-being | | | -0.504*** | | -0.43*** | | $F(4.57) = 47.037$,<br>$p < .001$ | 0.767 | 0.751 |

*Note.* N = 62. Standardized $\beta$ coefficients are displayed. All multiple regression analyses were controlled for age and sex. PSQI = Pittsburgh Sleep Quality Index, RIS = Regensburg Insomnia Scale, BDI = Beck Depression Inventory-II, AES = Apathy Evaluation Scale, FSMC = Fatigue Scale for Motor and Cognitive Functions, NMSQ = Non-Motor Symptoms Questionnaire.
\* $p \leq .05$ ; \*\* $p \leq .01$ ; \*\*\* $p \leq .001$ .

## 4 Discussion

This study identified reduced hrQoL in patients with iRBD independent of emerging PD motor symptoms. In particular, hrQoL was lower in patients with iRBD than HC in the SF-36 total score and the domains general health, vitality, and psychological well-being. In line with the affected hrQoL domains, diminished hrQoL in patients with iRBD was primarily correlated with more self-reported neuropsychiatric symptoms, sleep-wake disturbances, and autonomic symptoms. Multiple regression analyses revealed that significant factors linked to poorer hrQoL in patients with iRBD were more severe fatigue and depressive symptoms.

In patients with PD, sleep-wake disorders, including insomnia and excessive daytime sleepiness, are prevalent comorbidities closely related to poorer hrQoL^35^. Similarly, fatigue is reported by approximately half of the patients with PD^36,35,37^ and is associated with an increased α-synuclein oligomer level in the cerebrospinal fluid^38^. Lintel et al.^39^ argue that symptoms of fatigue overlap with the diagnostic criteria for depression and hence challenging to separate. This thinking aligns with our results that identify these two symptoms as the most significant factors associated with poorer hrQoL in patients with iRBD. Patients with iRBD reported more autonomic symptoms than HC but fewer than patients with PD. This finding indicates that non-motor symptoms, including autonomic complaints, may precede motor symptoms in emerging PD^11,40^. Other studies in PD patients support our results showing a pronounced link between non-motor symptoms and reduced hrQoL^13,14,35^.

Psychological well-being emerges as the most influential factor affecting self-reported hrQoL in prodromal PD. Depressive symptoms, in particular, have also been shown to be the most substantial contributor to hrQoL in patients with PD and are a well-known feature of prodromal PD^41,42,12^. Hence, clinicians should be highly attentive to these symptoms when assessing patients with (prodromal) PD. In line with our study, motor symptoms are less associated with poor hrQoL in patients with PD^14^, affirming the importance of non-motor symptoms for hrQoL in (prodromal) PD, irrespective of motor symptoms.

Strikingly, non-motor symptoms of patients with iRBD and PD are often overlooked and rarely treated in the clinical routine^43,44^. A study by Shulman et al.^44^ revealed that physicians were least sensitive to symptoms of fatigue in patients with PD. As a result, this clinically relevant symptom is at risk of being neglected. It is, however, essential to assess non-motor symptoms as this study underpins the impact on hrQoL in early disease stages. A positive correlation between non-motor symptom severity and a physician’s diagnostic sensitivity^44^ implies that patients with iRBD in the prodromal phase of PD might not receive treatment for their symptoms as they might still be mild. However, adequate treatment of non-motor symptoms is crucial due to the close link to increased functional disability, cognitive impairment, caregiver burden, and poor subjective hrQoL^45,46^.

Careful assessment of non-motor symptoms in patients with iRBD may also open the opportunity for treatment options for patients in the context of a missing disease-modifying therapy. This aspect is of utmost importance in the context of ethical considerations regarding an early diagnosis of PD – especially in its prodromal stage^47–49^. In a retrospective survey, most patients with PD were skeptical about early risk disclosure given missing pharmacological options^50^. However, a recent study by Pérez-Carbonell and colleagues demonstrated that most patients with iRBD appreciated an honest risk disclosure at the time of iRBD diagnosis^51^. Patients with early PD, who remained untreated, showed a decline of hrQoL over the next 18 months, whereas patients with initiated treatment showed a trend towards better hrQoL^52^. Together with our study linking reduced hrQoL to treatable neuropsychiatric non-motor symptoms in prodromal PD, this disease phase should not be solely regarded as a phase of watchful waiting but point physicians to provide adequate and prompt medical care to restore hrQoL.

A limitation of our study is the lack of longitudinal data. Additionally, the study was conducted in recently diagnosed “*de novo”* patients with iRBD. As a result, the self-reported depressive symptoms may have been aggravated due to the recent diagnosis^16,53^. Also, sample sizes regarding the PD and HC participants were relatively small. Still, a considerable advantage of the current study is the possibility to compare HC, patients with iRBD, and patients with PD to assess hrQoL across healthy aging, prodromal and manifest PD. Another strength of the present study is the well-characterized sample due to the comprehensive clinical assessments. Our results warrant further analysis of hrQoL in iRBD in larger data sets.

In summary, in clinical practice, careful screening for non-motor symptoms in patients with iRBD is crucial as present non-motor complaints might complicate diagnosis and treatment. Depressive symptoms, particularly fatigue, are the most relevant therapeutic targets in the prodromal stage of PD to enhance patients’ quality of life. Various treatment options for non-motor symptoms are available and their use is likely to increase patients’ well-being.

## Acknowledgments

We thank all study participants for their participation.

## Authors’ Roles

(1) Research project: A. Conception, B. Organization, C. Execution; (2) Statistical analysis: A. Design, B. Execution, C. Review and critique; (3) Manuscript: A. Writing of the first draft, B. Review and critique.

S.R.: 2A, 2B, 3A

M.-S. L.: 1B, 1C, 3B

A.S.: 1B, 1C, 3B

J.K.: 1B, 1C, 3B

K.-L.W.: 2C, 3B

C.E.J.D.: 1A, 1B, 1C, 2C, 3B

G.R.F.: 1A, 2C, 3B

A.O.: 2A, 2B, 3B

M.S.: 1A, 1B, 1C, 2A, 2C, 3A

Financial Disclosures of all authors (for the preceding 12 months)

S.R., M.-S.L., A.S., J.K. and K.-L.W. report no disclosures. C.E.J.D. received grants from the Clinician Scientist Program (CCSP), funded by the German Research Foundation (DFG, FI 773/15-1). G.R.F. receives royalties from the publication of the books Funktionelle MRT in Psychiatrie und Neurologie, Neurologische Differentialdiagnose, and SOP Neurologie and received honoraria for speaking engagements from Forum für medizinische Fortbildung FomF GmbH as well as grants from Deutsche Forschungsgemeinschaft (DFG, German Research Foundation), Project-ID 431549029, SFB 1451. A.O. received a grant from the Koeln Fortune Program (grant-no. 329/2021), Faculty of Medicine, University of Cologne, and the “Novartis-Stiftung für therapeutische Forschung”, both outside the submitted work. M.S. received grants from the Else Kröner-Fresenius-Stiftung (grant number 2019_EKES.02), and the Koeln Fortune Program, Faculty of Medicine, University of Cologne. M.S. receives funding from the program “Netzwerke 2021”, an initiative of the Ministry of Culture and Science of the State of Northrhine Westphalia. The sole responsibility for the content of this publication lies with the authors.

## Funding agencies

S.R., M.-S.L., A.S., J.K., and K.-L.W. report no disclosures. C.E.J.D. is supported by the Clinician Scientist Program (CCSP), funded by the Deutsche Forschungsgemeinschaft (DFG, German Research Foundation, FI 773/15-1). G.R.F. receives royalties from the publication of the books Funktionelle MRT in Psychiatrie und Neurologie, Neurologische Differentialdiagnose, and SOP Neurologie and received honoraria for speaking engagements from Forum für medizinische Fortbildung FomF GmbH as well as grants from Deutsche Forschungsgemeinschaft (DFG, German Research Foundation), Project-ID 431549029, SFB 1451. A.O. received a grant from the Koeln Fortune Program (grant-no. 329/2021), Faculty of Medicine, University of Cologne, and the “Novartis-Stiftung für therapeutische Forschung”, both outside the submitted work. M.S. received grants from the Else Kröner-Fresenius-Stiftung (grant number 2019_EKES.02), and the Koeln Fortune Program, Faculty of Medicine, University of Cologne. M.S. receives additional funding from the program “Netzwerke 2021”, an initiative of the Ministry of Culture and Science of the State of Northrhine Westphalia. The sole responsibility for the content of this publication lies with the authors.

## References

1. Sateia MJ. International Classification of Sleep Disorders-Third Edition. Chest. 2014;146(5):1387–1394. doi:10.1378/chest.14-0970

2. Schenck CH, Högl B, Videnovic A, eds. Rapid-Eye-Movement Sleep Behavior Disorder. Springer International Publishing; 2019. doi:10.1007/978-3-319-90152-7

3. Atik A, Stewart T, Zhang J. Alpha-Synuclein as a Biomarker for Parkinson’s Disease: Alpha-Synuclein as a Biomarker for PD. Brain Pathol. 2016;26(3):410–418. doi:10.1111/bpa.12370

4. Siderowf A, Concha-Marambio L, Lafontant DE, et al. Assessment of heterogeneity among participants in the Parkinson’s Progression Markers Initiative cohort using α-synuclein seed amplification: a cross-sectional study. Lancet Neurol. 2023;22(5):407–417. doi:10.1016/S1474-4422(23)00109-6

5. Stefani A, Iranzo A, Holzknecht E, et al. Alpha-synuclein seeds in olfactory mucosa of patients with isolated REM sleep behaviour disorder. Brain. 2021;144(4):1118–1126. doi:10.1093/brain/awab005

6. Iranzo A, Fernández-Arcos A, Tolosa E, et al. Neurodegenerative Disorder Risk in Idiopathic REM Sleep Behavior Disorder: Study in 174 Patients. Toft M, ed. PLoS ONE. 2014;9(2):e89741. doi:10.1371/journal.pone.0089741

7. Berg D, Postuma RB, Adler CH, et al. MDS research criteria for prodromal Parkinson’s disease: MDS Criteria for Prodromal PD. Mov Disord. 2015;30(12):1600–1611. doi:10.1002/mds.26431

8. Cochen De Cock V, Dotov D, Lacombe S, et al. Classifying Idiopathic Rapid Eye Movement Sleep Behavior Disorder, Controls, and Mild Parkinson’s Disease Using Gait Parameters. Mov Disord. 2022;37(4):842–846. doi:10.1002/mds.28894

9. Ehgoetz Martens KA, Matar E, Hall JM, et al. Subtle gait and balance impairments occur in idiopathic rapid eye movement sleep behavior disorder. Mov Disord. 2019;34(9):1374–1380. doi:10.1002/mds.27780

10. Postuma RB, Lang AE, Gagnon JF, Pelletier A, Montplaisir JY. How does parkinsonism start? Prodromal parkinsonism motor changes in idiopathic REM sleep behaviour disorder. Brain. 2012;135(6):1860–1870. doi:10.1093/brain/aws093

11. Aguirre-Mardones C, Iranzo A, Vilas D, et al. Prevalence and timeline of nonmotor symptoms in idiopathic rapid eye movement sleep behavior disorder. J Neurol. 2015;262(6):1568–1578. doi:10.1007/s00415-015-7742-3

12. Sumi Y, Masuda F, Kadotani H, Ozeki Y. The prevalence of depression in isolated/idiopathic rapid eye movement sleep behavior disorder: A systematic review and meta-analysis. Sleep Med Rev. 2022;65:101684. doi:10.1016/j.smrv.2022.101684

13. Soh SE, Morris ME, McGinley JL. Determinants of health-related quality of life in Parkinson’s disease: A systematic review. Parkinsonism Relat Disord. 2011;17(1):1–9. doi:10.1016/j.parkreldis.2010.08.012

14. Ophey A, Eggers C, Dano R, Timmermann L, Kalbe E. Health-Related Quality of Life Subdomains in Patients with Parkinson’s Disease: The Role of Gender. Park Dis. 2018;2018:1–9. doi:10.1155/2018/6532320

15. Kim KT, Motamedi GK, Cho YW. Quality of life in patients with an idiopathic rapid eye movement sleep behaviour disorder in Korea. J Sleep Res. 2017;26(4):422–427. doi:10.1111/jsr.12486

16. Seger A, Ophey A, Heitzmann W, et al. Evaluation of a Structured Screening Assessment to Detect Isolated Rapid Eye Movement Sleep Behavior Disorder. Mov Disord. Published online April 18, 2023:mds.29389. doi:10.1002/mds.29389

17. Nasreddine ZS, Phillips NA, Bédirian V, et al. The Montreal Cognitive Assessment, MoCA: A Brief Screening Tool For Mild Cognitive Impairment. J Am Geriatr Soc. 2005;53(4):695–699. doi:10.1111/j.1532-5415.2005.53221.x

18. Postuma RB, Berg D, Stern M, et al. MDS clinical diagnostic criteria for Parkinson’s disease: MDS-PD Clinical Diagnostic Criteria. Mov Disord. 2015;30(12):1591–1601. doi:10.1002/mds.26424

19. Beck AT, Steer RA, Brown G. Beck Depression Inventory–II. Published online 1996. doi:10.1037/t00742-000

20. Ware JE, Sherbourne CD. The MOS 36-item short-form health survey (SF-36). I. Conceptual framework and item selection. Med Care. 1992;30(6):473–483.

21. Buysse DJ, Reynolds CF, Monk TH, Berman SR, Kupfer DJ. The Pittsburgh sleep quality index: A new instrument for psychiatric practice and research. Psychiatry Res. 1989;28(2):193–213. doi:10.1016/0165-1781(89)90047-4

22. Crönlein T, Langguth B, Popp R, et al. Regensburg Insomnia Scale (RIS): a new short rating scale for the assessment of psychological symptoms and sleep in insomnia; Study design: development and validation of a new short self-rating scale in a sample of 218 patients suffering from insomnia and 94 healthy controls. Health Qual Life Outcomes. 2013;11(1):65. doi:10.1186/1477-7525-11-65

23. Johns MW. A New Method for Measuring Daytime Sleepiness: The Epworth Sleepiness Scale. Sleep. 1991;14(6):540–545. doi:10.1093/sleep/14.6.540

24. Beck AT, Epstein N, Brown G, Steer RA. An inventory for measuring clinical anxiety: Psychometric properties. J Consult Clin Psychol. 1988;56(6):893–897. doi:10.1037/0022-006X.56.6.893

25. Beck AT, Steer RA, Brown G. Beck Depression Inventory–II. Published online September 12, 2011. doi:10.1037/t00742-000

26. Marin RS, Biedrzycki RC, Firinciogullari S. Reliability and validity of the apathy evaluation scale. Psychiatry Res. 1991;38(2):143–162. doi:10.1016/0165-1781(91)90040-V

27. Penner I, Raselli C, Stöcklin M, Opwis K, Kappos L, Calabrese P. The Fatigue Scale for Motor and Cognitive Functions (FSMC): validation of a new instrument to assess multiple sclerosis-related fatigue. Mult Scler J. 2009;15(12):1509–1517. doi:10.1177/1352458509348519

28. Jessen F, Amariglio RE, Boxtel M, et al. A conceptual framework for research on subjective cognitive decline in preclinical Alzheimer’s disease. Alzheimers Dement. 2014;10(6):844–852. doi:10.1016/j.jalz.2014.01.001

29. Kalbe E, Bintener C, Ophey A, et al. Computerized Cognitive Training in Healthy Older Adults: Baseline Cognitive Level and Subjective Cognitive Concerns Predict Training Outcome. Health (N Y). 2018;10(01):20–55. doi:10.4236/health.2018.101003

30. Visser M, Marinus J, Stiggelbout AM, Van Hilten JJ. Assessment of autonomic dysfunction in Parkinson’s disease: The SCOPA-AUT. Mov Disord. 2004;19(11):1306–1312. doi:10.1002/mds.20153

31. Chaudhuri KR, Martinez-Martin P, Schapira AHV, et al. International multicenter pilot study of the first comprehensive self-completed nonmotor symptoms questionnaire for Parkinson’s disease: The NMSQuest study: Nonmotor Symptoms and PD. Mov Disord. 2006;21(7):916–923. doi:10.1002/mds.20844

32. Goetz CG, Tilley BC, Shaftman SR, et al. Movement Disorder Society-sponsored revision of the Unified Parkinson’s Disease Rating Scale (MDS-UPDRS): Scale presentation and clinimetric testing results: MDS-UPDRS: Clinimetric Assessment. Mov Disord. 2008;23(15):2129–2170. doi:10.1002/mds.22340

33. IBM SPSS Statistics for Windows. Published online 2021.

34. Posit team. RStudio: Integrated Development Environment for R. Published online 2022. http://www.posit.co/

35. Martinez-Martin P, Rodriguez-Blazquez C, Kurtis MM, Chaudhuri KR, on Behalf of the NMSS Validation Group. The impact of non-motor symptoms on health-related quality of life of patients with Parkinson’s disease. Mov Disord. 2011;26(3):399–406. doi:10.1002/mds.23462

36. Barone P, Antonini A, Colosimo C, et al. The PRIAMO study: A multicenter assessment of nonmotor symptoms and their impact on quality of life in Parkinson’s disease. Mov Disord. 2009;24(11):1641–1649. doi:10.1002/mds.22643

37. Siciliano M, Trojano L, Santangelo G, De Micco R, Tedeschi G, Tessitore A. Fatigue in Parkinson’s disease: A systematic review and meta-analysis: Fatigue in Parkinson’s Disease. Mov Disord. 2018;33(11):1712–1723. doi:10.1002/mds.27461

38. Zuo LJ, Yu SY, Wang F, et al. Parkinson’s Disease with Fatigue: Clinical Characteristics and Potential Mechanisms Relevant to α-Synuclein Oligomer. J Clin Neurol. 2016;12(2):172. doi:10.3988/jcn.2016.12.2.172

39. Lintel H, Corpuz T, Paracha S ur R, Grossberg GT. Mood Disorders and Anxiety in Parkinson’s Disease: Current Concepts. J Geriatr Psychiatry Neurol. 2021;34(4):280–288. doi:10.1177/08919887211018267

40. Durcan R, Wiblin L, Lawson RA, et al. Prevalence and duration of non-motor symptoms in prodromal Parkinson’s disease. Eur J Neurol. 2019;26(7):979–985. doi:10.1111/ene.13919

41. Leentjens AFG, Van den Akker M, Metsemakers JFM, Lousberg R, Verhey FRJ. Higher incidence of depression preceding the onset of Parkinson’s disease: A register study. Mov Disord. 2003;18(4):414–418. doi:10.1002/mds.10387

42. Schrag A. Quality of life and depression in Parkinson’s disease. J Neurol Sci. 2006;248(1-2):151–157. doi:10.1016/j.jns.2006.05.030

43. Baig F, Lawton M, Rolinski M, et al. Delineating nonmotor symptoms in early Parkinson’s disease and first-degree relatives. Mov Disord. 2015;30(13):1759–1766. doi:10.1002/mds.26281

44. Shulman LM, Taback RL, Rabinstein AA, Weiner WJ. Non-recognition of depression and other non-motor symptoms in Parkinson’s disease. Parkinsonism Relat Disord. 2002;8(3):193–197. doi:10.1016/S1353-8020(01)00015-3

45. Leroi I, McDonald K, Pantula H, Harbishettar V. Cognitive Impairment in Parkinson Disease: Impact on Quality of Life, Disability, and Caregiver Burden. J Geriatr Psychiatry Neurol. 2012;25(4):208–214. doi:10.1177/0891988712464823

46. Schrag A. What contributes to quality of life in patients with Parkinson’s disease? J Neurol Neurosurg Psychiatry. 2000;69(3):308–312. doi:10.1136/jnnp.69.3.308

47. Malkani RG, Wenger NS. REM Sleep Behavior Disorder as a Pathway to Dementia: If, When, How, What, and Why Should Physicians Disclose the Diagnosis and Risk for Dementia. Curr Sleep Med Rep. 2021;7(3):57–64. doi:10.1007/s40675-021-00206-1

48. Schaeffer E, Toedt I, Köhler S, Rogge A, Berg D. Risk Disclosure in Prodromal Parkinson’s Disease. Mov Disord. 2021;36(12):2833–2839. doi:10.1002/mds.28723

49. Vertrees S, Greenough GP. Ethical Considerations in REM Sleep Behavior Disorder: Contin Lifelong Learn Neurol. 2013;19(1):199–203. doi:10.1212/01.CON.0000427223.22963.d4

50. Schaeffer E, Rogge A, Nieding K, et al. Patients’ views on the ethical challenges of early Parkinson disease detection. Neurology. 2020;94(19):e2037–e2044. doi:10.1212/WNL.0000000000009400

51. Pérez-Carbonell L, Simonet C, Chohan H, et al. The Views of Patients with Isolated Rapid Eye Movement Sleep Behavior Disorder on Risk Disclosure. Mov Disord. Published online April 12, 2023:mds.29403. doi:10.1002/mds.29403

52. Grosset D, Taurah L, Burn DJ, et al. A multicentre longitudinal observational study of changes in self reported health status in people with Parkinson’s disease left untreated at diagnosis. J Neurol Neurosurg Psychiatry. 2006;78(5):465–469. doi:10.1136/jnnp.2006.098327

53. Paulsen JS, Nance M, Kim JI, et al. A review of quality of life after predictive testing for and earlier identification of neurodegenerative diseases. Prog Neurobiol. 2013;110:2–28. doi:10.1016/j.pneurobio.2013.08.003

